# Developing Patient-Reported Experience Measures (PREMs) and Patient-Reported Outcome Measures (PROMs) for Maternity Care in Australia: A Study Protocol

**DOI:** 10.1101/2025.06.18.25329896

**Authors:** Lauren C Spark, Karen Schlage, Emily J Callander, Emily Leefhelm, Michelle B Hobday, Negash Wakgari, Zoe Bradfield

## Abstract

**Background:** With some of the lowest global perinatal mortality rates, Australia is one of the safest countries to give birth. Despite impressive outcomes in clinical care, the prevalence of birth trauma, mistreatment and disrespect in maternity care has been the subject of much scrutiny. There are no nationally standardized reporting measures to indicate whether care provided during the critical and transformative period of childbearing meets the health and wellbeing needs of those giving birth. Recent reviews of maternity Patient-Reported Experience Measures (PREMs) and Patient-Reported Outcome Measures (PROMs) worldwide indicate current tools are of poor quality and lack consumer co-design. This protocol describes the first phase of the Measuring what Matters to Australian Mothers (MMAMs) study, to develop consumer co-designed PREMs and PROMs for maternity care in Australia.

**Methods:** A four-step multi-methods approach will be adopted for tool development. (i) A rapid literature review and thematic synthesis of existing maternity PREMs and PROMs tools will inform mapping of domains, dimensions and items across the maternity care continuum to develop a conceptual survey framework. (ii) Multiple rounds of iterative consultation with expert maternity care consumers and maternity healthcare and policy experts will refine the conceptual framework. (iii) A three-round modified e-Delphi study will be undertaken with a minimum n=50 maternity care consumers and n=50 health professionals to identify the priority items for inclusion in a maternity care PREM and PROM. (iv) Cognitive focus group interviews with maternity care consumers will assess the PREM and PROM tools for overall relevance, comprehensibility, and comprehensiveness. This approach will result in a draft set of PREMs and PROMs that will be piloted and validated across a nationally representative sample of maternity care consumers in Australia in phase two of the MMAMs study.

**Discussion:** This study will develop world-first consumer co-designed maternity care PREMs and PROMs. Following tool development outlined here, piloting, psychometric validation, and implementation planning phases will be undertaken. These tools will provide practical, brief and broad indicators of health quality, enabling benchmarking and improvements in the value and quality of person-centered maternity care at an individual, service and system level across Australia.

## Background

The World Health Organization recommends a universal approach to maternal health, emphasizing respectful and woman-centered care during pregnancy, childbirth, and the postnatal period (1). With some of the lowest global perinatal mortality rates, Australia is one of the safest countries in the world to give birth (2, 3). Despite impressive outcomes in clinical care, the prevalence of birth trauma, mistreatment and disrespect in maternity care have been the subject of much scrutiny (4–8).

Women’s experiences of maternity care impact a range of perinatal health outcomes (9). Negative experiences during childbirth can influence feelings of powerlessness, embarrassment, guilt, fear, perceptions of pain and development of more serious presentations such as trauma or post-traumatic stress disorder (10). By comparison, positive experiences in childbirth can influence functional mother-infant bonding, improved adaptation and transition to the new parenting role, and perceptions about future childbirth experiences (11). Measuring care experiences is necessary to improve the quality of care and, ultimately, care outcomes. Globally, patient-reported care experiences and outcomes are measured using standardized tools such as Patient-Reported Experience Measurements (PREMs) and Patient-Reported Outcome Measurements (PROMs) (12–16).

PREMs and PROMs have been used to evaluate healthcare quality standards in general populations worldwide for at least the last 30 years (17). PREMs provide a means to measure individuals’ experiences of health care, evaluating what happened during a care encounter and how this was received from the perspective of the service user (14, 18). By contrast, PROMs enable measurement of individuals’ appraisal of their outcomes, or, ‘what condition they are left in’ after a health care encounter in relation to their health, including physical, psychological and social functioning, and symptom severity (18, 19).

Studies have demonstrated the role of PREMs and PROMs to improve the safety and quality of care, reducing over or under-servicing, contributing to higher-value care that meets population needs, and increasingly as a metric for health service accreditation and funding models (17, 20, 21). However, PREMs and PROMs in maternity care are a relatively contemporary phenomenon. Two recent systematic reviews synthesized the validity of 25 maternity PREMs (14) and nine PROMs (19) around the world. Authors found most had poor quality evidence underpinning instrument development, with scant description of how women were involved in tool development, and little detail regarding evidence of content validity (14, 19).

Whilst previous studies have advanced our understanding of what women want from maternity care in Australia (22–25), the current safety and quality benchmarking of Australian maternity care focuses exclusively on clinical outcomes. This approach provides a narrow understanding of trends and what aspects of maternity care need to be improved. There are no nationally standardized PREMs and PROMs to indicate whether care provided during the transformative period of childbearing meets the full health and wellbeing needs as identified by those giving birth. The Australian National Maternity Strategy (26) identified the need for a national standard and measure of maternity PREMs and PROMs as an area for priority investment, and several peak organizations and national studies have echoed this call for action (27–30).

The significant knowledge gap in evaluating and benchmarking the experiences and outcomes of maternity care in Australia from the perspective of the service user needs to be addressed to improve the safety and quality of maternity care. PREMs and PROMs offer a mechanism to ensure patient voices are central to ensuring healthcare is equitable, safe, patient-centered and high-quality (16). To meaningfully inform service improvements, patient-reported measures must be rigorously developed and designed in alignment with what women want from maternity care (14). This requires fundamental commitment to, and engagement in, co-design principles with maternity care consumers, healthcare professionals and policymakers to balance identified needs and a pragmatic approach to implementation to unlock the full potential of PREMs and PROMs to improve the value of care.

The aim of the Measuring what Matters to Australian Mothers (MMAMs) study is to develop, validate and plan implementation of national consumer co-designed maternity PREM and PROM safety and quality tools that best measure the experiences and outcomes recognized as most important by those giving birth. This protocol describes the first phase of the MMAMs study, to develop consumer co-designed PREMs and PROMs for maternity care in Australia. The resulting draft set of PREMs and PROMs will be piloted and validated across a nationally representative sample of maternity care consumers in Australia in the next phase of the MMAMs study.

## Methods

### Study design

A four-step multi-methods approach will be adopted for tool development: (i) A rapid literature review (31) and thematic synthesis (32, 33) of existing maternity care PREMs and PROMs will be conducted to map domains, dimensions and items to develop a draft conceptual survey framework; (ii) Multiple rounds of iterative consultation with n=11 expert maternity care consumers and n=14 maternity care health professional experts will refine the conceptual framework; (iii) A three-round modified e-Delphi study will be undertaken with a minimum n=50 maternity care consumers and n=50 health professionals to identify the priority items for inclusion in a maternity care PREM and PROM; (iv) Cognitive focus group interviews with up to n=15 maternity care consumers will assess the PREM and PROM tools for overall relevance, comprehensibility, and comprehensiveness.

### Approach

This development process has been informed and guided by COSMIN (COnsensus-based Standards for the selection of health Measurement INstruments), an international range of frameworks, guidelines and checklists to help guide the design, development and evaluation of PROMs (34–36). Although COSMIN is focused on PROMs, their frameworks have been used to evaluate the quality of both PREMs (14) and PROMs (19) in the maternity care space. COSMIN frameworks have been used to guide the current study design to ensure an evidence-based methodology to develop high-quality patient-reported tools that are relevant, comprehensible and comprehensive to maternity care consumers, whilst ensuring their value and impact as safety and quality tools to meaningfully influence system improvement.

### Governance

The MMAMs study will be led by a Curtin University Investigator Team (co-authors ZB, LS, KS, EC, EL), with project governance supported by three overarching expert advisory groups: (i) an independently chaired Executive Advisory Group (EAG) including representation (n=9) from pivotal peak health organizations and Commonwealth departments to provide high-level strategic guidance, policy advice and clinical expertise; (ii) an Expert Consumer Group (ECG) collectively representing a diverse sample of maternity care consumers, defined as aged over 18 years and had a baby in the Australian healthcare system within the last 10 years, with varied maternity care experiences to share the consumer voice and advocate consumer perspectives (n=11), led by a Curtin University-appointed Consumer Fellow (co-author KS); and, (iii) a Working Advisory Group (WAG) including members (n=5) who have national and/or jurisdictional responsibilities for operationalization of maternity care/safety and quality, with critical understanding of implementation of patient-reported measures to provide expert clinical, technical, operational and implementation advice. The representation and size of these groups will evolve as the study progresses. These groups, governed by Terms of Reference, will meet regularly throughout the project to provide advice and guidance, and will be referred to throughout the protocol as they will play a fundamental role in tool development.

### Step 1: Rapid literature review

A rapid literature review and thematic synthesis of existing maternity care PREMs and PROMs will be conducted to map domains, dimensions and items across the maternity care continuum to develop a draft conceptual survey framework. A compendium of relevant tools have been identified in recent systematic reviews of maternity PREMs (n=25) (14) and PROMs (n=9) (19) and will be used as the basis for this review. Citation tracking and hand searching will complement this search approach. Domains (broad general areas), dimensions (specific aspects within a domain) and items (survey questions) from existing measures will be mapped separately for: (i) experiences and (ii) outcomes; and by maternity care timepoints examined across the maternity care continuum: (i) antenatal, and/or (ii) intrapartum (labor and/or birth), and/or (iii) postpartum.

A thematic synthesis of this information will identify the domains and dimensions shared or unique to each timepoint, along with example item/s for each dimension. These findings will inform the development of a draft conceptual framework that summarizes the key elements assessed in existing maternity PREMs and PROMs. The draft framework will be reviewed independently and then collectively by members of the Investigator Team, with the aim to reduce duplication, remove ambiguity, and simplify framework elements. Any edits or revisions will be made by consensus amongst the Investigator Team.

### Step 2: Expert consultation

Multiple iterative consultations over a four-month period will be conducted with expert maternity consumers (ECG members n=11) and expert maternity care health professionals, policy and system experts (EAG members n=9 and WAG members n=5) during formal regular project group meetings to gather feedback and advice to further refine the conceptual framework. These conversations will be guided by semi-structured questions and focus on the exploration of priority and additional survey elements to measure maternity care experiences and outcomes at specific timepoints, in addition to discussion on the importance (relevance), clarity, and comprehensiveness of draft framework elements. Group meetings will be recorded and transcribed. After each meeting, the Investigator Team will critically review, analyze and refine the draft conceptual framework as needed. This process will result in a draft set of PREMs and PROMs in preparation for the round one e-Delphi survey.

### Step 3: Delphi study

#### Approach

A three-round modified e-Delphi study will be undertaken to identify the priority items for inclusion in a maternity care PREM and PROM. The e-Delphi technique is a well-established online method that uses a structured, systematic process involving iterative rounds of communication, facilitation and controlled feedback to elicit consensus among anonymous expert panel members on a specific topic. The e-Delphi technique is a time and cost-effective approach to obtaining expert group opinion and insight, whereby collective judgements are valued over individual perspectives and inherent bias like dominance and group conformity are eliminated (37).

The e-Delphi will utilize an online method of survey distribution and completion for each round, hosted on the Qualtrics survey platform (Qualtrics, Provo, UT). One survey link will be generated for each survey round. Each round is anticipated to assess PREM content followed by PROM content. There may be multiple versions of each PREM and PROM to assess, depending on how many timepoints are determined to be needed (i.e., different items may be relevant to antenatal vs intrapartum vs postpartum timepoints). Each survey round is anticipated to take about 30 minutes to complete.

#### Eligibility

To be eligible to participate, all participants must satisfy specific expert selection criteria as either a consumer or professional. For consumers, this is defined as: (i) aged 18 years and over, and (ii) had a baby within the Australian maternity system in the last 10 years. For professionals, this is defined as having professional experience in any of: (i) maternity care (clinical, leadership, policy, service delivery); and/or (ii) safety and quality benchmarking/assessment/monitoring specific to health service delivery; and/or (iii) patient reported measure development/implementation/evaluation. Due to the professional and personal demands on consumers and other experts, it is not feasible to require participants to participate in each of the three survey rounds. Therefore, participants are not required to complete previous rounds to be eligible to participate in subsequent rounds.

#### Sample size

COSMIN guidelines indicate the sample size of a quantitative (survey) study used to identify relevant content for a patient-reported measure should be large enough to assume that saturation was reached (36). In line with recommendations to achieve a ‘very good’ rating, a sample size of a minimum of 100 participants is forecasted to participate in Round 1 of this study (36). This is estimated to include a minimum n=50 maternity care consumers, and n=50 professionals.

#### Recruitment

The recruitment process will adopt intentional approaches to engage with an accessible and diverse potential participant pool, combining snowball recruitment and purposive recruitment for maximum variation. Combining these recruitment methods can be beneficial in achieving both diversity and depth of engagement (38).

Recruitment e-flyers will be developed for each round to refer interested participants directly to the Qualtrics survey via a hyperlink. Invited survey participants will include: (i) members of the EAG, WAG and ECG; (ii) other individuals and groups of relevant consumers and health professionals as identified by these group members; and (iii) other individuals and groups selected intentionally to represent specific characteristics and perspectives, monitored and determined ad-hoc based on the demographic responses of participants who have completed the survey to date.

Prior to Round 1, the Investigator Team will liaise with EAG, WAG and ECG group members to collectively develop a Master Recruitment List of expert contacts. This discussion will be guided by the aim of ensuring broad representation and participation amongst diverse consumer and health professional groups. Members may be tasked with disseminating the flyer to their relevant contacts at the commencement of each round, as agreed, to leverage their existing networks and influence. Participants will also have the opportunity to leave their email address at the end of each survey round to receive an email inviting them to participate in subsequent rounds. Each round will be open for an estimated minimum of two weeks, guided by response rates. Depending on the response rate, up to two email prompt reminders may be distributed to identified groups or individuals during this time.

Participant demographics will be monitored during the survey open period. Should there be an identified lack of participation from key groups or populations (e.g., midwives; consumers living in rural and remote areas), members of the Investigator Team and the EAG, EAG and ECG may be called upon to reach out to their specific target networks to share the recruitment flyer with the aim to increase participation amongst these identified groups.

#### Process

At the commencement of each round within Qualtrics, participants will view: (i) a Participant Information Statement providing an overview of the study; (ii) PREMs and PROMs Briefing Information about the purpose of tool development; (iii) Summarized feedback from previous rounds (for Rounds 2 and 3); (iv) a Participant Consent Statement to confirm understanding and provide consent; (v) demographic questions to complete; and (vi) current round of survey questions to complete.

## Round 1

### Data collection

Each PREMs and PROMs survey will provide a list of potential survey items. Items will be listed within each domain, and dimensions for each item will be provided in brackets [e.g., PREM Domain 1: Communication; Item 1: I was given enough of the right information to make informed decisions about my care (decision-making)]. Participants will be asked to: (i) rate each item according to how important (relevant) and how clear, relevant and feasible it is for a health service to use results from this item to make meaningful service improvements (actionable), using a four-point scale (1 = not important or not actionable – could exclude; 2 = somewhat important and may be actionable; 3 = quite important and actionable; 4 = very important and actionable – must include); and (ii) provide open-response feedback to identify concerns or suggestions with respect to the importance (relevance), completeness (comprehensive), and clarity (comprehensibility) of each item, in addition to where there may be any gaps or duplication of content.

### Data analysis

De-identified data will be analyzed descriptively using SAS 9.4 (SAS Institute Inc.). Within each round, data will be analyzed separately for each PREM and PROM survey. The distribution of responses across each of the four-point scale items will be reported. To identify which items will be retained for Round 2, responses will be coded either as ‘not or somewhat important and not or somewhat actionable’ (0) or ‘quite or very important and actionable’ (1). A total score will then be calculated for each item by summing the scores of all participants divided by the total number of participants in the Round 1 sample, to identify the proportion of agreement overall. In the absence of a formal guideline, the threshold for inclusion and exclusion of items will be set at 75% (0.75) agreement (i.e., consensus threshold of three quarters or more participants): (i) Items that score ≥0.75 without any suggested revision required will be retained; (ii) Items that score ≥0.75 with suggested revision will be considered and revised by the Investigator Team based on consensus agreement; (iii) Items that score <0.75 without comment will be dropped; (iv) Items that score <0.75 with suggested revision will be considered and revised by the Investigator Team based on consensus agreement.

A sub-analysis will compare responses between consumers and health professionals. This will be done by summing the scores of: (i) all consumers divided by the total number of consumers in the Round 1 sample; and (ii) all professionals divided by the total number of professionals in the Round 1 sample (proportion of agreement by group). If the difference in agreement between groups is ≥0.25 (25%), the Investigator Team will report where this disagreement lies. Items that do not meet the overall consensus threshold of 75% agreement, but do meet the threshold based upon consumer responses only, will be considered to be included in the subsequent round by the Investigator Team based on consensus agreement. This will be done to ensure consumer input is considered in isolation of health professional perspectives and maintain importance of consumer co-design in tool development.

Content analysis of the qualitative feedback will be conducted using Microsoft Excel and/or NVivo (guided by the scope and complexity of feedback), reviewed independently and then collectively by the Investigator Team. Any edits will be agreed by consensus. These suggestions will be carefully considered so as not to undermine the e-Delphi consensus process itself (i.e., consideration of ‘minor’ vs ‘major’ revisions based on their potential significance and impact).

Summary results and feedback regarding participation, results and content changes will be provided to participants via email and made available on the Qualtrics landing page at the commencement of the next Round.

## Round 2

### Data collection

Items will be listed for each PREM and PROM survey as per Round 1. Participants will be asked to: (i) rank items in priority order of importance and actionability within each domain; and (ii) provide open-response feedback as per Round 1.

### Data analysis

Descriptive statistics reporting ranking frequency data will be calculated for each item in SAS: mean (range), median (interquartile range – IQR) and mode. This will result in a sequential ranking of items by importance within each domain, based upon median ranking scores. This will be completed for (i) all participants and (ii) separately for consumer and health professional groups. Ranking of the consumer and health professional groups will be compared for agreement using an appropriate statistical test such as the Kendall’s W (Kendall’s coefficient of concordance). The Investigator Team will independently and then collectively discuss findings. The number of items retained within each domain will be informed by the statistics and any clear threshold indicators across and/or within groups, as agreed by consensus. A content analysis approach and subsequent provision of feedback will follow the same as Round 1.

## Round 3

### Data collection

For each PREM and PROM survey, the topmost ranked items as identified from Round 2 analysis will be presented with the domain and dimension for each in brackets. Participants will be asked to: (i) rank items in priority order of importance and actionability, regardless of each domain; and (ii) provide open-response feedback as per previous rounds.

### Data analysis

Descriptive statistics will be calculated, sequentially ranked, and compared across and within groups as per Round 2. Similarly, findings will be discussed amongst the Investigator Team and retained items will be informed by the statistics and any clear threshold indicators across and/or within groups. A content analysis approach will follow as previous. This will result in a draft set of final items for each PREM and PROM survey.

### Step 4: Cognitive focus group interviews with consumers

#### Approach

Patient-reported measurement tools must be clear, simple, relevant, appropriate, comprehensive and easy to understand (36, 39–41). In line with COSMIN quality standards for the development of a patient-reported measure (36), a cognitive interview study or other pilot test should be performed with the target population to test the PROM for comprehensibility and comprehensiveness. The aim of the cognitive focus group interviews is to explore these concepts with maternity care consumers. Participants will be required to: (i) complete the draft PREM and PROM surveys online; then, (ii) participate in a small online focus group interview to provide their feedback. Participants will receive AUD$200 honorarium payment in total for participation in both survey and interview elements.

#### Eligibility

To be eligible to participate, consumers must be: (i) aged 18 years and over, and (ii) had a baby within the Australian maternity system in the last 10 years. Participants must at least partially complete the online survey to be eligible to participate in the group interview.

#### Sample size

COSMIN guidelines (36) indicate in a qualitative (interview) study where each item is tested in an appropriate number of consumers, a sample size of ≥ 7 is given a rating a ‘very good’ rating. In line with these recommendations, a minimum n=7 participants and maximum n=15 participants will be scheduled across 1-2 group interview timeslots.

#### Recruitment

Recruitment e-flyers will be developed to provide information on study eligibility and refer to the two study elements: (i) 15-20-minute online survey; and (ii) small online two-hour group interview. Interested individuals are encouraged to email the Project Manager to register their interest. The Project Manager will confirm eligibility, study details, and interview times with interested candidates on a first-come-first-served basis until at least the minimum n=7 spots have been allocated.

In the first instance, all current members of the ECG (n=11) will be emailed the recruitment e-flyer. If unable to obtain the minimum sample size, the Investigator Team will: (i) email consumers who participated in the e-Delphi study the recruitment e-flyer; and/or (ii) engage with other existing consumer connections to share a recruitment e-flyer to their consumer networks.

## Process

### Online survey

An estimated 21 days prior to the scheduled cognitive interview session, participants will receive a link to the PREM and PROM surveys hosted on Qualtrics (Qualtrics, Provo, UT), to be completed independently and unaided. Participants will view: (i) a Participant Information Statement providing an overview of the study; (ii) PREMs and PROMs Briefing Information about the purpose of the surveys and interviews; (iii) Participant Consent Statement to confirm understanding and provide consent for both the survey and interview components; (v) demographic questions to complete; and (vi) draft survey items.

Participants will be asked to follow the instructions to complete the PREM and PROM survey sequentially, rating each item as ‘easy’, ‘neutral’ or ‘difficult’, and encouraged to make notes next to each item to enhance clarity and understanding. At the end of the survey, participants will provide their email address and submit their responses. Respondents will be emailed a copy of their responses for their reference during the cognitive interview session. The survey must be completed within seven days of the scheduled interview.

### Data analysis

Online survey data will be analyzed descriptively using SAS 9.4 (SAS Institute Inc.). A content analysis of any data from free text fields will be conducted independently and then collectively by members of the Investigator Team to inform interview discussions.

### Cognitive focus group interviews

The Consumer Fellow (KS), skilled in consumer group facilitation and moderation, will facilitate the cognitive group interviews. The Consumer Fellow will refer to a semi-structured guide to discuss each item within each survey in turn. Participants will be prompted using a combination of verbal probing and think-aloud strategies (42, 43) to encourage participants to verbalize their approach in answering questions and enable the interviewer to query a participant’s understanding and interpretation.

Comprehensibility will be assessed to check the clarity, understanding, and language used specific to: (i) survey instructions; (ii) items; (iii) response options; and (iv) recall period. Comprehensiveness will be assessed to explore face validity and content validity to check: (i) all items are relevant, suitable and appropriate; (ii) whether the items together comprehensively cover the constructs the survey intends to measure; (iii) that all essential issues and concepts have been included; and (iv) if any important issues or concepts have been omitted.

Interviews will be recorded and transcribed. Following the session, the Consumer Fellow will provide a written reflection and brief notes.

### Data analysis

The interview transcripts and Consumer Fellow reflection and notes will be reviewed independently and then collectively by members of the Investigator Team to identify key themes and flag suggested ‘minor’ or ‘major’ changes within each survey. The analysis may result in a series of recommended changes to the survey. Members of the Investigator Team will meet to discuss findings, issues and recommended changes, and agree by consensus to changes required. Minor adjustments to the survey will require no further action. If major adjustments are needed, further engagement with EAG, WAG and/or ECG members will follow until the Investigator Team reach agreed consensus that all issues have been adequately addressed. This will result in the final PREMs and PROMs ready for large-scale national piloting to explore psychometric properties including validity and reliability.

## Discussion

This protocol describes the planned research approach to develop consumer co-designed maternity PREMs and PROMs tools to measure women’s experiences and maternity care outcomes in Australia. Equipping systems to meet the needs, preferences and outcomes identified by consumers is central to the value-based health care agenda (44). To date, women’s voices have been absent from determining what constitutes high-value pregnancy, birth and postpartum care. Understanding which experiences and outcomes ‘matter most’ to women and birthing people is critical to ensure meaningful improvements to the lives of those who give birth in Australia.

Existing global maternity PREMs and PROMs lack descriptions of women’s involvement with tool development, with insufficient descriptions of content validity (14, 19). Without measurable indicators to reflect the real-world experiences and outcomes reported by women, services remain restricted to using narrow, clinically defined indicators that provide an incomplete picture of safety and quality in maternity care (45). Aligned with value-based health care principles, the mission of the MMAMs study is to reorientate services and systems towards assessing the value of care against the experiences and outcomes identified to be pragmatic priorities by both maternity care consumers and healthcare experts.

This protocol describes the first phase of the MMAMs study, to develop consumer co-designed PREMs and PROMs to improve the quality and safety of maternity care and maternal health outcomes across Australia. The resulting draft set of PREMs and PROMs will be piloted and validated across a nationally representative sample of maternity care consumers in Australia in the next phase of the MMAMs study. Across these two phases, establishing adequate content validity that meaningfully involves women in determining what is relevant, comprehensive and comprehensible will be crucial to improving the evidence-base of maternity-specific patient-reported measures to inform value-based health service improvement (19). Whilst simultaneously progressed in parallel with all study phases, a formal implementation planning phase will constitute the final phase of the MMAMs study. Implementation planning will provide a framework of recommendations for the national scale-up of maternity PREMs and PROMs. This will involve extensive collaboration with services and organizations in Australia who have the agency, authority and remit to embed and roll out national maternity PREMs and PROMs.

Overall, this work has the potential to impact and improve the safety and quality of maternity care, to optimize women’s experiences and achieve clinical outcomes that meet the identified needs and priorities of women giving birth in Australia. The availability and use of PREMs and PROMs data derived from evidence-based, psychometrically-sound, consumer co-designed tools have the potential to facilitate national benchmarking of maternity care, drive continuous system improvement, enhance patient safety and quality, and ultimately improve maternal health and wellbeing.

## Declarations

### Ethics approval and consent to participate

This study has been approved by the Curtin University Human Research Ethics Committee (HRE2025-0200) and will be carried out in line with the Australian National Health and Medical Research Council (NHMRC) National Statement in Ethical Conduct in Human Research (2023). Informed consent will be obtained from participants.

### Consent for publication

Not applicable.

### Availability of data and materials

There is no primary data available for this protocol.

### Authors’ contributions

LS led the conceptualization and development of the operational study protocol that provided the foundation for this manuscript, led development and review of this manuscript, and is the MMAMs Project Manager. KS, EC and EL are members of the Investigator Team and provided critical input into the operational study protocol, development and revision of this manuscript. MH supported elements of data extraction for literature review and synthesis, and provided review and revision of this manuscript, particularly the approach to data analysis. NW supported the development, preparation and review of this manuscript for submission. ZB conceptualized and designed this study, provided critical guidance and input into the development and revision of the operational study protocol and this manuscript, and is the Principal Investigator of the broader MMAMs study. All authors contributed to the content of the article and approved the submitted version.

## Data Availability

There is no primary data available for this protocol.

## Acknowledgements

This project is made possible through the generous goodwill of our valued collaborators and stakeholders who see value and impact in partnering with us throughout this project to improve women’s health and the quality and safety of maternity care across Australia. The MMAMs Investigator Team warmly acknowledge Professor Caroline Homer for her foundational contributions related to the establishment of project governance and in her ongoing role as independent chair of the MMAMs EAG. We sincerely thank and acknowledge the valuable time and profound contributions of all our committed EAG, WAG and ECG members, and also extend a warm thank you to all future study participants.

## List of abbreviations

COSMIN: COnsensus-based Standards for the selection of health Measurement INstruments
EAG: Executive Advisory Group
ECG: Expert Consumer Group
MMAMs: Measuring what Matters to Australian Mothers
PREMs: Patient-Reported Experience Measures
PROMs: Patient-Reported Outcome Measures
SAS 9.4: Statistical Analysis System, version 9.4
WAG: Working Advisory Group

